# Long-term immunogenicity of BNT162b2 vaccination in the elderly and in younger health care workers

**DOI:** 10.1101/2021.08.26.21262468

**Authors:** Pinkus Tober-Lau, Tatjana Schwarz, Kanika Vanshylla, David Hillus, Henning Gruell, EICOV/COVIM Study Group, Norbert Suttorp, Irmgard Landgraf, Kai Kappert, Joachim Seybold, Christian Drosten, Florian Klein, Florian Kurth, Victor Max Corman, Leif Erik Sander

## Abstract

COVID-19 mRNA vaccine BNT162b2 is highly immunogenic and effective, but recent studies have indicated waning anti-SARS-CoV-2 immune responses over time. Increasing infection rates has led authorities in several countries to initiate booster campaigns for vulnerable populations, including the elderly. However, the durability of vaccine-induced immunity in the elderly is currently unknown. Here, we describe interim results of a prospective cohort study comparing immune responses in a cohort of vaccinated elderly persons to those in healthcare workers (HCW), measured six months after first immunisation with BNT162b2. Anti-SARS-CoV-2 S1-, full Spike- and RBD-IgG seropositivity rates and IgG levels at six months were significantly lower in the elderly compared to HCW. Serum neutralization of Delta VOC measured by pseudovirus neutralisation test was detectable in 43/71 (60.6%, 95%CI: 48.9-71.1) in the elderly cohort compared to 79/83 in the HCW cohort (95.2%, 95%CI: 88.3-98.1) at six months post vaccination. Consistent with the overall lower antibody levels, SARS-CoV-2-S1 T cell reactivity was reduced in the elderly compared to HCW (261.6 mIU/ml, IQR:141.5-828.6 vs 1198.0 mIU/ml, IQR: 593.9-2533.6, p<0.0001).

Collectively, these findings suggest that the established two-dose vaccination regimen elicits less durable immune responses in the elderly compared to young adults. Given the recent surge in hospitalisations, even in countries with high vaccination rates such as Israel, the current data may support booster vaccinations of the elderly. Further studies to determine long-term effectiveness of COVID-19 vaccines in high-risk populations and the safety and effectiveness of additional boosters are needed.

## Main text

COVID-19 mRNA vaccine BNT162b2 (Comirnaty) is highly immunogenic and effective against symptomatic SARS-CoV-2 infection.^1^ Recent studies have indicated decreasing anti-SARS-CoV-2 antibody levels, but largely stable vaccine efficacy six months after initial vaccination.^2,3^ The emergence of variants of concern (VOC), particularly Delta (B.1.617.2), has raised concerns about waning protection, particularly in risk populations such as the elderly, who display reduced immune responses to BNT162b2 vaccination compared to younger adults.^4^ In light of increasing infection rates caused by the Delta VOC, authorities in several countries, including in the United States, Israel, and Germany, are discussing or have recently begun offering a booster dose to vulnerable populations, such as all persons over 60 years of age. However, the durability of vaccine-induced immunity in the elderly is currently unknown. Following up on a previous analysis, we describe interim results of a prospective cohort study comparing immune responses in a cohort of vaccinated elderly persons to those in healthcare workers (HCW)^4,5^, measured six months after first immunisation with BNT162b2.

Six-month follow-up visits were completed by 107 HCW (median age 35 [IQR: 30-48] years, 60.7% female) and 82 elderly persons (median age 82.5 [IQR: 78-87] years, 74.4% female) (**Appendix Table 1**). Twelve participants were excluded from further analysis due to RT-PCR- or serologically confirmed SARS-CoV-2 infection at baseline or before the second vaccination (**Appendix Table 1**). Anti-SARS-CoV-2 S1-IgG seropositivity rates at six months were lower in the elderly (48/80, 60.0%, 95%CI: 49.0-70.0) compared to HCW (95/97, 97.9%, 95%CI: 92.8-99.6; p<0.0001). Median anti-SARS-CoV-2 S1-IgG levels were also lower in the elderly (1.2 S/Co, IQR: 0.5-2.2) than in HCW (median 3.2 S/Co, IQR: 2.4-4.1, p<0.0001) (**Fig 1A**). Similar results were obtained for serum anti-RBD- and anti-full spike-IgG levels (**Appendix Fig. S1B, C**). In line with reduced seropositivity and antibody levels, we observed significantly lower surrogate virus neutralization (sVNT) titres among the elderly (56.6%, IQR: 30.1-69.6) compared to HCW (88.1%, IQR: 79.3-93.1, p<0.0001) (**Appendix Fig. S1D**). Pseudovirus neutralization test (pNT) revealed a significant decline of serum neutralization of Delta VOC from two months after first vaccination (i.e. four weeks after second vaccination). Serum neutralization of Delta VOC was detectable in 43/71 (60.6%, 95%CI: 48.9-71.1) in the elderly cohort compared to 79/83 in the HCW cohort (95.2%, 95%CI: 88.3-98.1) at six months post vaccination (**Fig. 1B**). Similarly, neutralization of Alpha VOC was observed in 79/83 (95.2%, 95%CI: 88.3-98.1) HCW versus 49/71 (69.0%, 95%CI:57.5-78.6) of the elderly cohort six months after vaccination (**Fig. S1E**). Mean neutralizing titres for both Alpha and Delta VOCs were significantly lower in the elderly cohort (geometric mean ID_50_ Alpha: 20.15, 95%CI:15.34-26.46; Delta: 14.5, 95%CI: 11.5-18.2) compared to HCW (Alpha: 134.4, 95%CI:104.2-173.4; Delta: 72.7, 95%CI: 58.7-89.9, p<0.0001). The recent rise of viral variants carrying immune escape spike mutations like E484K, may further reduce immunity in the elderly. Binding capacity of serum antibodies to RBDs with distinct mutation patterns (K417N/T, L425R, T478K, E484K/Q, N501Y, and E484Q) found in six known SARS-CoV-2 variants were significantly lower in the elderly compared to the HCW cohort (p<0.0001 for all RBDs, **Appendix Fig. S1F**). Consistent with the antibody responses, SARS-CoV-2-S1 T cell reactivity was reduced in the elderly compared to HCW (261.6 mIU/ml, IQR:141.5-828.6 *vs* 1198.0 mIU/ml, IQR: 593.9-2533.6, p<0.0001) (**Fig. 1C**).

**Figure 1:**
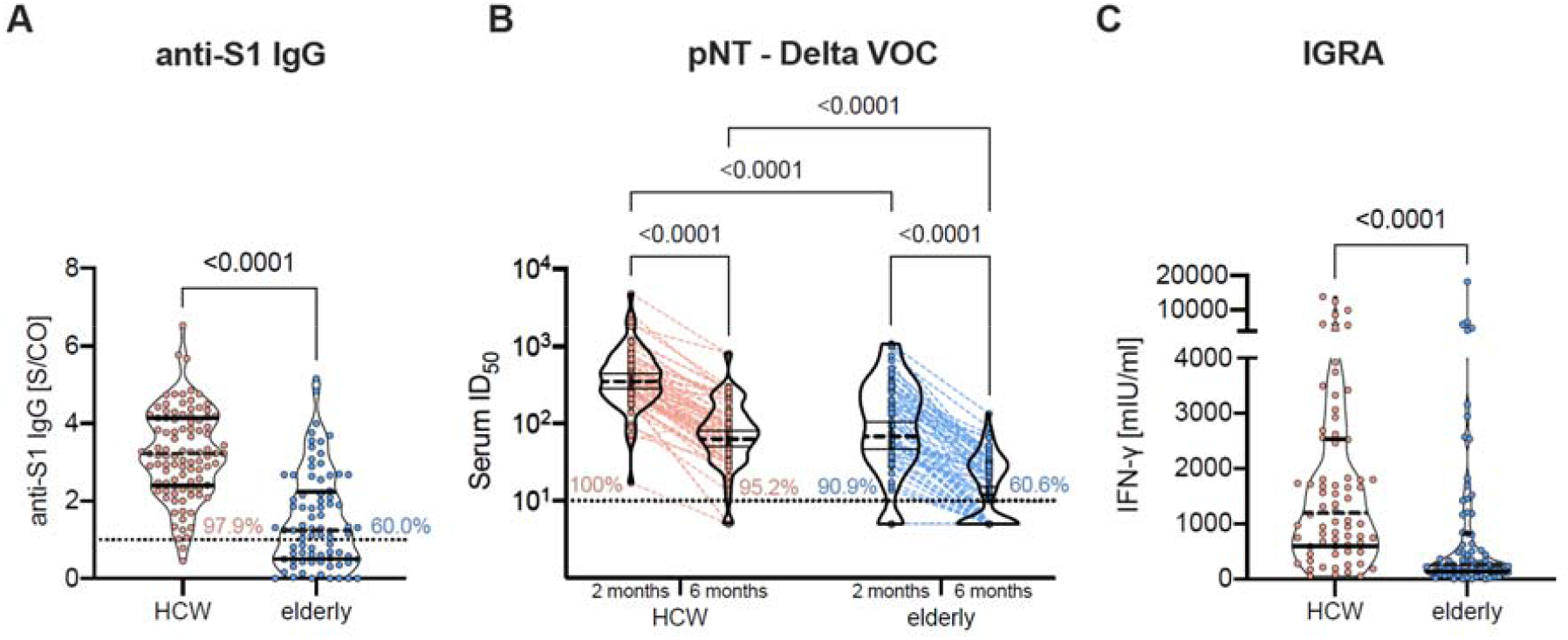
BNT162b2 induced SARS-CoV-2 antibody and T cell response six months after vaccination in HCW and elderly persons. **(A)** Anti-S1 IgG in serum measured by a microarray-based immunoassay, **(B)** serum pseudovirus neutralization against the Delta VOC two and six months after vaccination measured by pNT, and **(C)** SARS-CoV-2 S1 specific T cell response detected by IGRA. Dotted horizontal lines indicate the manufacturer’s threshold for anti-S1 IgG ≥1 S/Co (A) and the lower limit of detection (1:10 dilution) for pNT (B). Horizontal lines within plotted data regions indicate the median and interquartile range, except for pNT, where the geometric mean and 95% confidence interval is shown. P values (all <0.0001) were calculated by non-parametric Mann Whitney *U* test. S/Co: signal-to-cutoff, pNT: pseudovirus neutralization test, ID_50_: 50% inhibition dilution, IGRA: interferon-γ release assay, IU: international units.

The study provides evidence of waning immunity to COVID-19 vaccination in the elderly, with 39.4% of elderly participants lacking detectable serum neutralizing activity against the Delta variant six months after first vaccination with BNT162b2. Reassuringly, neutralizing capacity was still detectable in the vast majority of young adults. Decline in antibody levels in the first six months post vaccination is generally expected and has also been observed for COVID-19 vaccines.^3^ In our study, all investigated immunogenicity markers were markedly reduced in the elderly compared to younger HCW six months post vaccination. These data suggest that the established two-dose vaccination regimen elicits less durable immune responses in the elderly compared to young adults. Analyses of the phase 3 efficacy trial of BNT162b2 show that vaccine efficacy against symptomatic disease remains at 91% through a six-month follow-up.^2^ However, the Delta VOC was not prevalent during the observation period and the majority (59.3%) of the study participants were younger, aged 16-55 (median 51). Given the recent surge in cases and hospitalisations, even in countries with high vaccination rates like Israel, and evidence of increasing breakthrough infections, our data may support booster vaccinations of the elderly, given the strongly increased risk of severe disease and death with older age. Studies to determine long-term effectiveness of COVID-19 vaccines in high-risk populations are needed.

## Data Availability

Anonymised raw immunogenicity data for tested samples and anonymised reactogenicity data can be obtained upon request. Requests will be reviewed and approved by the authors, collaborators, and the security department on the basis of scientific merit and absence of competing interests.

## Acknowledgments

We thank all study participants at Charité – Universitätsmedizin Berlin for their participation. We also thank the entire staff of the Department for Occupational Medicine and the Charité Clinical Study Center at Charité – Universitätsmedizin Berlin and the Berlin Institute of Health for their support of the study. SARS-CoV-2 RBD variants antigens were kindly provided by InVivo BioTech Services GmbH (Hennigsdorf, Germany) to the Seramun Diagnostica GmbH (Heidesee, Germany; https://www.seramun.com).

## Appendix

**Figure S1:**
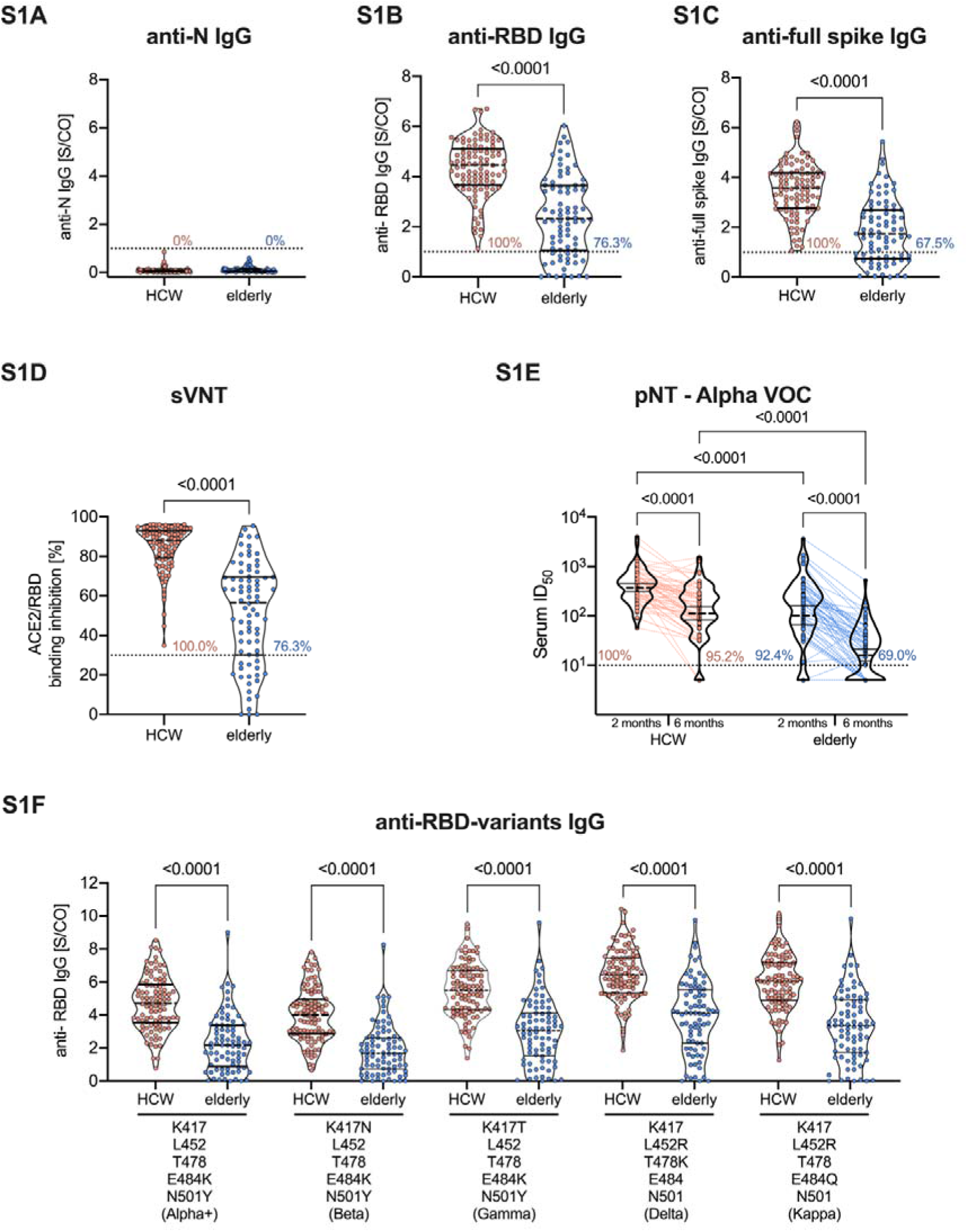
Anti-SARS-CoV-2 nucleocapsid protein, RBD, and full spike IgG response 6 months after vaccination. (**A**) Anti-SARS-CoV-2 N, (**B**) RBD- (**C**) and full-spike IgG measured in the serum of BNT162b2 vaccinated HCW and elderly persons six months after the first vaccination. (**D**) Neutralizing capacity was measured by sVNT and (**E**) serum neutralization against Alpha (B.1.1.7) VOC detected by pNT in vaccinated HCW and elderly persons six months after the first vaccination. (**F**) Binding capacity of serum IgG against six different RBDs of SARS-CoV-2 variants carrying the indicated mutations in HCW and elderly, measured by ELISA. Dotted lines indicate the manufacturer’s threshold values: for anti-N, anti-RBD, and anti-full spike IgG ≥1 S/Co, for sVNT >30%, and the lower limit of detection (1:10 dilution) for pNT. Lines indicate the median and interquartile range except for pNT, where the geometric mean and 95% confidence interval are shown. P values were calculated by the non-parametric Mann Whitney *U* test or Kruskal-Wallis test with Dunn’s multiple comparisons test. S/Co: signal-to-cutoff, N: nucleocapsid protein, RBD: receptor-binding domain, sVNT: surrogate virus neutralization test, ACE2: angiotensin-converting enzyme 2, ID50: 50% inhibition dilution.

**Table 1.**
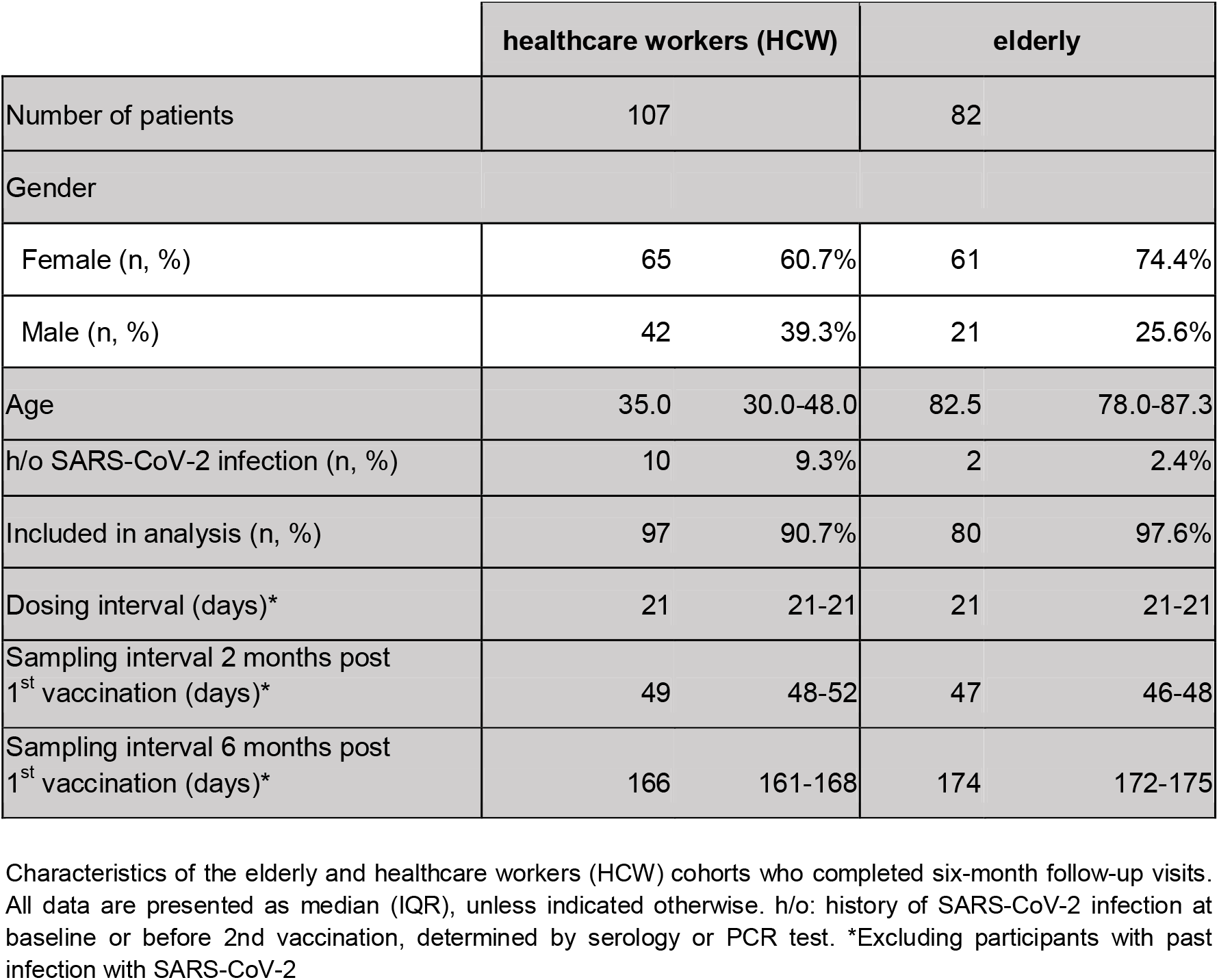

### Study participants

Study participants were recruited in the EICOV, COVIMMUNIZE, and COVIM studies, three prospective cohort studies conducted under the auspices of Charité - Universitätsmedizin Berlin, Germany, in accordance with the Declaration of Helsinki and Good Clinical Practice.^4,5^ EICOV and COVIMMUNIZE studies were approved by the local ethics committee of Charité - Universitätsmedizin Berlin (EA4/244/20, EA4/245/20), and COVIM was approved by the Federal Institute for Vaccines and Biomedicines (Paul Ehrlich Institute) and by the ethics committee of the state of Berlin (EudraCT-2021–001512–28). Written informed consent was obtained by all participants, according to local and national regulations. Blood sampling was conducted before 1st vaccination, 4±1 weeks after the 1st and 2nd vaccinations, respectively, and 6±1 months after the 1st vaccination. SARS-CoV-2 reverse transcription PCR (RT-PCR) of oropharyngeal swabs were performed during study visits in order to detect concomitant infections. Study participants were eligible for analysis of immunogenicity if they had completed a course of two vaccinations with BNT162b2 and the first dose was administered at least 22 weeks before the 6-month visit.

### Antibody assessment and Interferon-γ Release of SARS-CoV-2–Specific T Cells

Blood samples were tested for anti-SARS-CoV-2 antibodies, neutralizing capacity, and T cell reactivity as previously described^5^. In brief, SARS-CoV-2 specific antibodies were quantified using the commercially available SeraSpot^®^ Anti-SARS-CoV-2 IgG microarray-based immunoassay including nucleocapsid and spike as antigens (Seramun Diagnostica GmbH, https://www.seramun.com), allowing for differentiation between infection and vaccine-induced immune responses. Functional neutralization capacity was investigated using a commercially available ELISA-based SARS-CoV-2 RBD-ACE2 binding inhibition assay (surrogate SARS-CoV-2 neutralization test (sVNT) cPass (medac GmbH, https://international.medac.de). A lentivirus-based SARS-CoV-2 pseudovirus neutralization assay (pNT) was employed to determine serum 50% inhibitory dilutions (ID_50_) against Alpha (B.1.1.7) and Delta (B.1.617.2) variants of concern.^6^ SARS-CoV-2 specific T cell responses were measured by an interferon-□ release assay (IGRA) of S1 stimulated T cells in whole blood using a commercially available kit (EUROIMMUN AG, https://www.euroimmun.de). SARS-CoV-2 RBD variants antibody testing was performed using RBD proteins (provided by InVivo BioTech Services GmbH, Hennigsdorf, Germany). Different RBD proteins were printed in an array format in a similar manner as done for the screening SeraSpot® Anti-SARS-CoV-2 IgG microarray (Seramun Diagnostica GmbH, https://www.seramun.com) and tested the same way.

